# Baseline epigenetics as a biomarker of mepolizumab response in severe asthma

**DOI:** 10.1101/2025.09.02.25334098

**Authors:** Hongmei Zhang, Xin Jin, Hasan Arshad, Ramesh J Kurukulaaratchy

## Abstract

Biologic treatment options have expanded considerably for severe asthma, transforming patient management, but only ~70% of patients respond to the treatment following standard criteria.

This preliminary study utilized data collected in the Wessex AsThma CoHort of difficult asthma (WATCH) study. DNAm was measured in n=15 severe asthma patients at baseline, i.e., before mepolizumab (MEPO), an interleukin-5 receptor antagonist, was administered and the response to MEPO was analyzed. We aimed to assess the potential of DNA methylation as a feasible biomarker to predict responses to MEPO among severe asthma patients.

Our preliminary findings suggested that pre-biologic DNA methylation as a predictor of a patient’s response status may outperform pre-biologic blood eosinophils cell counts.

## Introduction

Over the last decade, biologic treatment options have expanded considerably for severe asthma, transforming patient management.^1^ Real-world studies have shown that while biologic treatments are effective, only ~70% of patients respond to the treatment following standard criteria.^2^ Interest has focused on pre-biologic clinical measures such as blood eosinophil count [BEC], total IgE, exhaled nitric oxide [FeNO], asthma control or exacerbation rate to predict biologic response^3^ and guide treatment selection. However, the predictive quality and real-world utilization of those markers are unclear,^4^ resulting in delays in receiving effective treatment, ongoing exacerbation-risk and exposures to harmful therapies like oral corticosteroids [OCS] while healthcare providers face the high costs of unsuccessful biologic trials.^5^ There is therefore a need to “get biologic selection right first time” in severe asthma.

It has been shown that both childhood and adult asthma, along with its symptoms, are associated with epigenetic modifications, e.g., DNA methylation (DNAm).^6^ The potential of DNAm as a biomarker to predict monoclonal antibody (biologic) drug responsiveness has been reported in other health conditions such as rheumatoid arthritis^7^ and male idiopathic infertility^8^. Success in other fields suggests a possibility of pre-treatment DNAm as a predictive marker for biologic responsiveness in severe asthma.

## Methods

This preliminary study utilized data collected in the Wessex AsThma CoHort of difficult asthma (WATCH) study. DNAm was measured in n=15 severe asthma patients at baseline, i.e., before mepolizumab (MEPO), an interleukin-5 receptor antagonist, was administered and the response to MEPO was analyzed. We aimed to assess the potential of DNA methylation as a feasible biomarker to predict responses to MEPO among severe asthma patients. Details of the WATCH study, DNA methylation assessment and rigorous methods of CpG sites screening and statistical analyses are in the Online Repository.

## Results

Biologic response was defined as a 50% reduction in asthma exacerbations needing OCS or in maintenance OCS dose without loss of asthma control after 12 months MEPO.^9^ Among the 15 severe asthma patients, 7 were males, of whom 4 (57.1%) were responsive to MEPO. For the 8 females, 5 (62.5%) were responsive.

After quality control, 692,432 CpGs underwent multi-layer screening (Figure 1a, Figure S1 in Online Repository), which resulted in 12 CpGs with each as a candidate predictor. Variations were observed in means and SDs in DNAm across the 12 CpGs as well as between responsiveness and non-responsiveness (Figure 1b). Using 5-fold cross-validation (with data of 12 subjects as training and 3 as testing), we evaluated each of the 12 CpGs’ predictability based on logistic regression with Firth’s bias reduction due to small sample size. Across all the 12 CpGs, averages of area under the curves (AUCs) across the five sets of training samples ranged from 0.949 to 1.0. Average sensitivities for the five set of validation samples across the 12 CpGs were between 0.800 and 1, and average specificities were between 0.875 and 1.0 (Online Repository Table S1). The sensitivity and specificity were determined at a threshold such that the sum of the two was maximized.

**Figure 1.**
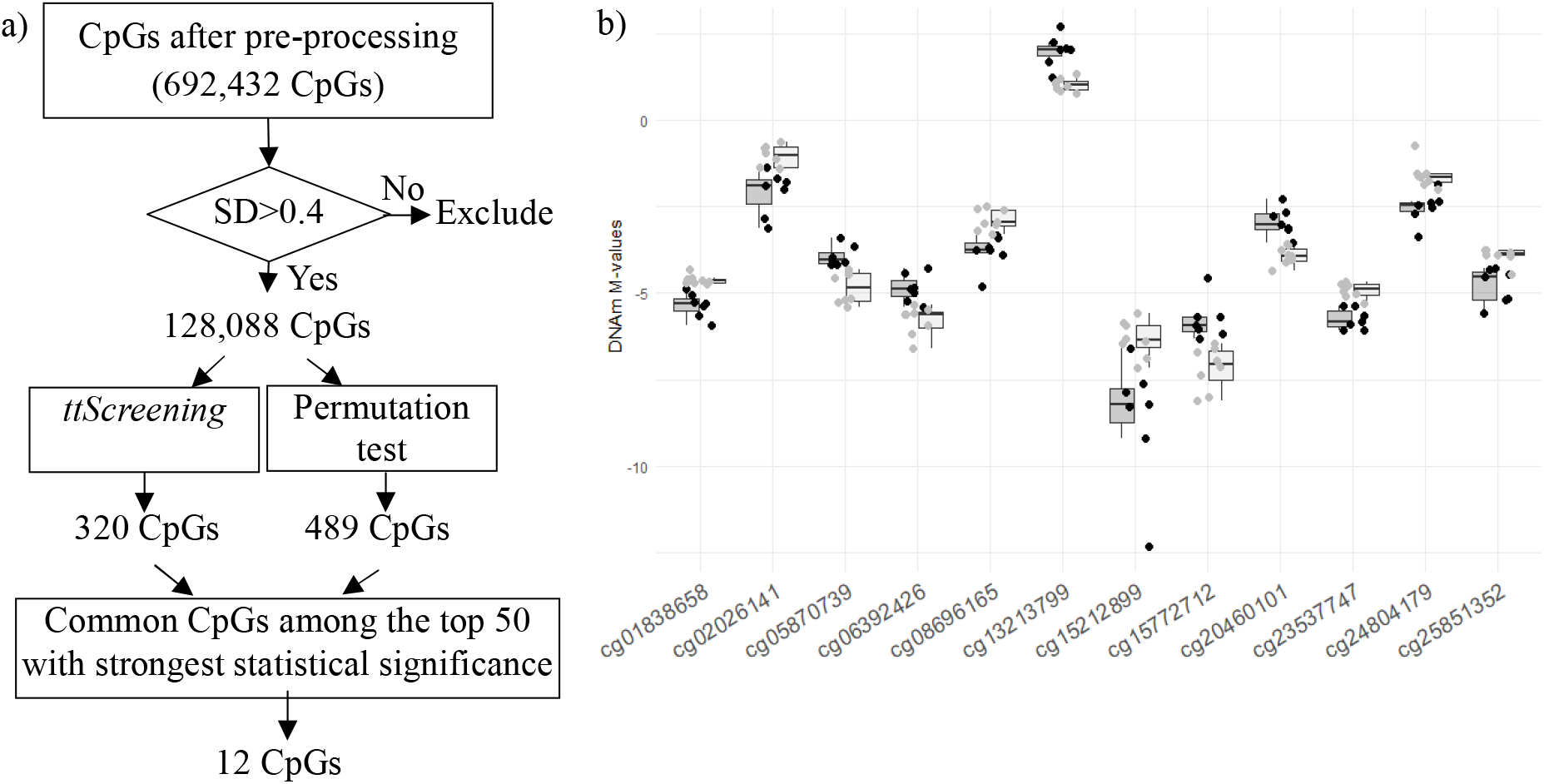
a) Flow chart of the multi-layer screening. SD: standard deviation. Threshold 0.4 was determined by the pattern of scree plot following the empirical elbow rule (Figure S1in Online Repository). b) Boxplots of DNA methylation at the 12 candidate CpGs. X-axis: CpGs sites; Y-axis: DNA methylation in M-values. Light gray box and dots: responders; Dark gray box and black dots: non-responders.

As MEPO was selected based on pre-treatment BEC, we also evaluated BEC’s predictability using the same approach as that for CpGs. The average AUC from the training samples was 0.702, average sensitivity and specificity for the validation samples were both 0.6. When using all the data, the AUC was 0.705 with a sensitivity of 0.625 and specificity of 0.714. Given that MEPO was selected based on BEC, the statistics reported here, although all were substantially lower than those based on CpGs, were already biased toward high values. Our preliminary findings suggested that pre-biologic DNA methylation as a predictor of a patient’s response status may outperform pre-biologic BEC.

## Discussion

Since the sample size is small, even though we implemented techniques including multi-layer screening and cross-validation to improve the validity of the identified markers, there is a possibility that the markers detected in our study are false positives. To address this limitation, in addition to the need of a large-scale study, replication of the findings in independent cohorts is crucial to the quality and generalizability of identified markers. We also wish to highlight that, since genes work jointly, addressing joint activities among CpGs while identifying predicting markers is necessary to diminish the risk of biased and incomplete predictive markers.

In summary, we currently lack markers that can reliably predict biologic response in severe asthma. Although replicating the findings in a large and independent cohort is warranted, these 12 CpGs are promising candidate predictors for the outcome of reducing asthma exacerbations needing OCS; a key indicator for prescribing biologics. Asthma has multiple impacts and the value of these CpGs towards MEPO response to other asthma outcomes like asthma control, lung function and quality of life warrant further attention. In addition, the association of these (and other) CpGs with response to other biologic treatments is a key future research need to guide accurate biologic treatment selection.

## Summary

- Currently we lack markers that can reliably predict biologic response in severe asthma.
- Pre-biologic DNA methylation as predictor of a patient’s response to MEPO may outperform clinical markers.

## Online Repository

https://osf.io/zq9ws/files/osfstorage, which is also available in the Supplemental files.

## Data Availability

The data that support the findings of this study are available on request from the corresponding author. The data are not publicly available due to privacy or ethical restrictions.

## Abbreviations

AUC: Area under the curve
BEC: Blood eosinophil cell
CpG: Cytosine-phosphate-guanine
DNAm: DNA methylation
FeNO: Exhaled nitric oxide
MEPO: Mepolizumab
OCS: Oral corticosteroids
WATCH: Wessex AsThma CoHort of difficult asthma

## Acknowledgements

The authors highly appreciated the participation of severe asthma patients in this WATCH study and are thankful to the High-Performance Computing facility at the University of Memphis (UofM) that helped tremendously with the detection of epigenetic markers and to Chitra Ghosh at UofM for her valuable input. HZ’s work was partially supported by R21AI175891 related to study design and analyses of DNA methylation data.

## REFERENCES

1. Brusselle GG, Koppelman GH. Biologic Therapies for Severe Asthma. The New England journal of medicine. Jan 13 2022;386(2):157–171. doi:10.1056/NEJMra2032506

2. Fong WCG, Azim A, Knight D, et al. Real-world Omalizumab and Mepolizumab treated difficult asthma phenotypes and their clinical outcomes. Clinical and experimental allergy : journal of the British Society for Allergy and Clinical Immunology. Aug 2021;51(8):1019–1032. doi:10.1111/cea.13882

3. Scelo G, Tran TN, L. TT, et al. Exploring Definitions and Predictors of Response to Biologics for Severe Asthma. J Allergy Clin Immunol Pract. Sep 2024;12(9):2347–2361. doi:10.1016/j.jaip.2024.05.016

4. Albers FC, Licskai C, Chanez P, et al. Baseline blood eosinophil count as a predictor of treatment response to the licensed dose of mepolizumab in severe eosinophilic asthma. Respiratory medicine. Nov 2019;159:105806. doi:10.1016/j.rmed.2019.105806

5. Reibman J, Tan L, Ambrose C, et al. Clinical and economic burden of severe asthma among US patients treated with biologic therapies. Annals of Allergy, Asthma & Immunology. 2021;127(3):318–325.e2.

6. Sheikhpour M, Maleki M, Ebrahimi Vargoorani M, Amiri V. A review of epigenetic changes in asthma: methylation and acetylation. Clin Epigenetics. Mar 29 2021;13(1):65. doi:10.1186/s13148-021-01049-x

7. Plant D, Wilson AG, Barton A. Genetic and epigenetic predictors of responsiveness to treatment in RA. Nat Rev Rheumatol. Jun 2014;10(6):329–37. doi:10.1038/nrrheum.2014.16

8. Lujan S, Caroppo E, Niederberger C, et al. Sperm DNA Methylation Epimutation Biomarkers for Male Infertility and FSH Therapeutic Responsiveness. Sci Rep. Nov 14 2019;9(1):16786. doi:10.1038/s41598-019-52903-1

9. Health NIf, Excellence C. Mepolizumab for treating severe refractory eosinophilic asthma. NICE London; 2017.

