## Supplemental material for "Baseline epigenetics as a biomarker of mepolizumab response in severe asthma"

The following material is also available at the Online Repository: https://osf.io/zq9ws/files/osfstorage

**SUPPLEMENTAL MATERIAL FOR ONLINE REPOSITORY**

**METHODS**

**The study population**

WATCH (n= 500; ClinicalTrials.gov ID NCT03996590) is an ongoing longitudinal clinical observational cohort study of difficult to treat asthma based at University Hospitals Southampton NHS Foundation Trust (UHSFT), Southampton, United Kingdom (UK), established in 2015 and approved by West Midlands – Solihull Research Ethics Committee (REC reference: 14/WM/1226). A detailed outline of WATCH study methodology has been published.^1^ Patients managed with British Thoracic Society Step “high dose therapies” and/or “continuous or frequent use of oral corticosteroids” in the Adult and/or Transitional Regional Asthma Clinic at UHSFT were invited to participate following informed consent. Data acquisition at enrolment included detailed clinical, health and disease-related questionnaires, anthropometry, allergy skin prick testing (SPT), spirometry fractional exhaled nitric oxide (FeNO), clinically requested blood tests and collection of biological samples (blood and urine).

For this pilot study, data and samples were analysed from patients who were biologic naïve at WATCH enrolment but subsequently commenced MEPO. Blood eosinophil count (BEC) analysed in this study were the highest recorded values before biologics and within 6 months of WATCH study enrolment. Biologic response was defined in line with the National Institute for Clinical Excellence criteria as a 50% reduction in exacerbations or maintenance OCS dose without loss of asthma control after 12 months MEPO.^2^

**DNA methylation (DNAm)**

DNA was extracted from pre-biologic whole blood of 15 severe asthma patients via a standard salting out procedure^3^. Following the manufacturer’s standard protocol, one microgram DNA was bisulfite-treated using the EZ 96-DNA methylation kit (Zymo Research, CA, USA) for cytosine to thymine conversion. Epigenome-wide DNAm was measured by the Illumina Infinium MethylationEPIC BeadChip (>850,000 CpGs) (Illumina, Inc., San Diego, CA, USA). CpGs that did not achieve a detection p-value of 10^−16^ in at least 95% of samples were excluded, as well as CpGs with probe SNPs within 10 base pairs and with a minor allele frequency > 0.007. We also excluded methylation sites on sex chromosomes to avoid potential bias. After pre-processing, 692,432 CpG sites were included in the subsequent statistical analyses. Intensities of DNAm were quantile normalized using the R computing package, *minfi*. Methylation levels of each CpG site (β values) ranging from 0 (no cytosine methylation) to 1(complete cytosine methylation) were calculated as a ratio of methylated (Me) to the sum of methylated and unmethylated (U) probes (β = Me/(c + Me + U)), with a constant (c) to avoid division by zero. To reduce heteroscedasticity in β values, logit-transformed β-values of DNAm were used and noted as M values (*M = log2 (β/1 − β)*).

DNA methylation (DNAm) measured in blood can be influenced by variations in cell compositions. The goal of this study is to utilize DNAm to predict biologic treatment response and cell compositions are potentially informative to such predictions. Henceforth, we did not correct for cell type heterogeneity among the patients when predicting their responses to the treatment.

*Screening of CpG sites*

With the small sample size (n=15), effort to improving prediction power is essential. We implemented two layers of screening. In the first layer, we focused on the variations in DNAm determined by sample standard deviations (SD). We excluded CpG sites with DNAm showing small variations; in particular, if a CpG site with DNAm standard deviation larger than a selected threshold, then this site was deemed to pass the first layer of screening and included in the second layer of screening. Selection of the threshold was driven by the empirical elbow rule applied to a scree plot of standard deviations.

In the second layer of screening, we adopted two independent methods to screen in parallel to further exclude potentially uninformative CpGs. One approach was to use robust regressions with surrogate variables and training-testing technique implemented in the R package *ttScreening.*^4,5^ We adopted the default setting suggested by the *ttScreening* package except that we used one random split with half of the samples for training and the remaining for testing, instead of the default setting of 2/3 and 1/3, respectively, due to small sample size. CpGs showing statistical significance in both training and testing pair (statistical significance level was 0.05 and 0.1, respectively) were kept. The other approach was non-parametric permutation tests; we permuted the responses to biologic treatment to generate samples under the null and, for each permuted data, calculated mean difference in DNAm between responders and non-responders, based on which an empirical p-value for the original mean difference in DNAm at a given CpG site was calculated. In total, we 9,999 permutations were employed to estimate the statistical significance of each CpG site. A p-value < 0.05 was deemed as being statistically significant.

Finally, among the top 50 CpGs showing the strongest statistical significance and with p-value<0.05 from each of the two parallel screening, common CpGs between the two were included in the subsequent statistical analyses for prediction.

**Statistical analyses**

*Association between DNAm and response*

The goal was to examine the potential of predictability of the CpGs that passed screening with respect to a patient’ responses to the biologic treatment, MEPO. CpGs that passed the multi-layer screening were included in this analysis and logistic regressions were applied for this purpose,

*Logit* *(P(Response=Yes))=log (*$\frac{P (Response =Yes)}{1-P (Response =Yes)}$*) =*$\beta_{0}$+$\beta_{k}\times X_{k}$

In the expression above, *X_k_* refers to DNA methylation (in M-values) at a candidate CpG site *k*, *k=1*, …, *K* and $\beta_{k}$ is regression coefficient with $\beta_{k}\neq0$ indicating that CpG *k* is a potentially important predictor for response status. For a given candidate CpG, the estimated $\beta_{0}$ and $\beta_{k}$were used to calculate the probability of being responsive to the drug for each subject, based on which response status of that subject could be predicted.

In addition to DNA methylation, we also estimated the predictability of BEC. The same statistical model was applied with corresponding updated interpretation of *X_k_* and $\beta_{k}.$

*Statistics to assess a predictive model*

To address sampling error as well as small sample size, we implemented a five-fold cross-validation. In particular, for each model, we split the data into five groups with each group three subjects. For each fold, we used data in the 12 subjects (training data) to fit a regression model, based on which we inferred the area under the curve (AUC) and identified the threshold that maximizes the sum of sensitivity and specificity. The threshold was then used to determine the response status of the remaining three subjects (testing data). Finally, AUC, sensitivity, and specificity summarizing the current model were the mean of the five AUCs, five sensitivities, and five specificities, respectively. All the analyses were conducted in R^6^ computing packages and SAS^7^.

**Scree plot of the SDs**

The following plot is a scree plot of standard deviations (SDs) in DNAm (M values) at 692,432 CpGs that passed quality control. Turning point 0.4 was selected and used to exclude CpGs showing consistently small variations based on the assumption of sparsity of informative CpGs with respect to biologic drug treatment responsiveness. To be conservative, we took a relatively small cutoff 0.4.

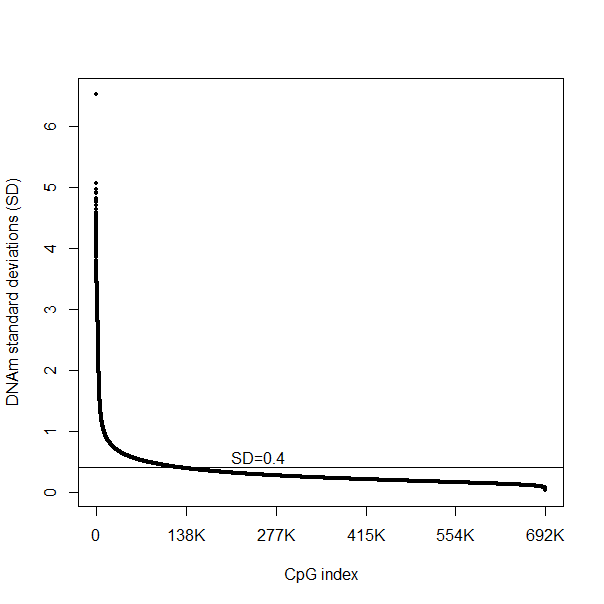

Figure S1. Scree plot of SD (DNA methylation in M values) at 692,432 CpGs.

**Predicting statistics of BEC and the candidate CpGs**

| Table S1. Predictive models and their corresponding prediction statistics (on averages from cross-validation samples) and identified thresholds (based on average thresholds from cross-validation samples). The threshold is in the scale of log-odds ratio. ECC: Eosinophil cell counts. | | | | | | | |
| --- | --- | --- | --- | --- | --- | --- | --- |
| Model | Variables | Gene | Location | AUC | Threshold | Sensitivity | Specificity |
| 1 | ECC |  |  | 0.702 | 0.242 | 0.600 | 0.600 |
| 2 | cg25851352 | *TRAPPC9* | Body | 0.949 | 0.325 | 0.933 | 1.000 |
| 3 | cg15212899 | *FBXO24;LRCH4* | TSS200; TSS200 | 0.966 | 0.096 | 0.800 | 0.875 |
| 4 | cg02026141 | *Intergenic* |  | 0.949 | -0.301 | 1.000 | 0.875 |
| 5 | cg06392426 | *KRI1* | Body | 0.983 | 0.333 | 0.933 | 0.875 |
| 6 | cg13213799 | *Intergenic* |  | 0.983 | 0.232 | 0.933 | 0.875 |
| 7 | cg24804179 | *PRPH* | Body | 0.983 | 0.070 | 0.800 | 0.875 |
| 8 | cg08696165 | *SIX3* | 3’UTR | 1.000 | -0.044 | 0.933 | 0.875 |
| 9 | cg01838658 | *Intergenic* |  | 1.000 | 0.531 | 1.000 | 1.000 |
| 10 | cg05870739 | *PCDHGA* | 5’UTR | 1.000 | -0.092 | 1.000 | 1.000 |
| 11 | cg15772712 | *TOB2* | TSS1500 | 1.000 | -0.038 | 1.000 | 1.000 |
| 12 | cg20460101 | *CDK6* | 5’UTR | 1.000 | 0.421 | 0.800 | 0.875 |
| 13 | cg23537747 | *PODN* | TSS200 | 1.000 | 0.256 | 0.900 | 1.000 |

**References**

1.	Azim A, Mistry H, Freeman A, et al. Protocol for the Wessex AsThma CoHort of difficult asthma (WATCH): a pragmatic real-life longitudinal study of difficult asthma in the clinic. *BMC Pulm Med*. May 24 2019;19(1):99. doi:10.1186/s12890-019-0862-2

2.	Health NIf, Excellence C. Mepolizumab for treating severe refractory eosinophilic asthma. NICE London; 2017.

3.	Miller SA, Dykes DD, Polesky HF. A simple salting out procedure for extracting DNA from human nucleated cells. *Nucleic Acids Res*. Feb 11 1988;16(3):1215. doi:10.1093/nar/16.3.1215

4.	*ttScreening: Genome-wide DNA methylation sites screening by use of training and testing samples (R Package)*. 2014. http://cran.fhcrc.org/web/packages/ttScreening/index.html

5.	Ray MA, Tong X, Lockett GA, Zhang H, Karmaus WJ. An Efficient Approach to Screening Epigenome-Wide Data. Research Support, N.I.H., Extramural. *Biomed Res Int*. 2016;2016:16. doi:10.1155/2016/2615348

6.	*R: A language and environment for statistical computing.* . 2018.

7.	*SAS/STAT Software, Version 9.4*. 2018.
